# Long-term impacts of Legionnaires’ disease on health and well-being: rationale, study design and baseline findings of a matched cohort study (LongLEGIO)

**DOI:** 10.1101/2024.08.20.24312294

**Authors:** Melina Bigler, Malina Vaucher, Manuel Wiederkehr, Sophia Brülisauer, Werner C. Albrich, Sarah Dräger, Valentin Gisler, Isabel Akers, Daniel Mäusezahl

## Abstract

**Background:** The long-term effects of Legionnaires’ disease beyond the acute infection and their impact on healthcare utilisation remain poorly understood. We present the rationale and study design of a matched prospective observational cohort study (*LongLEGIO*) aimed at investigating the persistent sequelae on patients’ health, well-being, and health service use following community-acquired Legionnaires’ disease, compared to other bacterial pneumonias that tested negative for *Legionella*.

**Methods:** Patients with Legionnaires’ disease and other bacterial *Legionella* test-negative pneumonia are recruited from secondary and tertiary care hospitals and matched for sex, age, hospital-level and date of diagnosis. Semi-structured interviews were conducted at baseline (shortly after the pneumonia diagnosis) and at two, six and 12 months following appropriate antibiotic therapy. Baseline assessments capture pre-existing conditions, illness experience, and disease severity, while follow-up assessments evaluate long-term symptoms, healthcare utilisation, quality of life (EQ-5D-5L), and social/work impacts. Data on case management and the disease severity are extracted from patient records.

**Results:** A total of 59 patients with community-acquired Legionnaires’ disease and 60 patients with other bacterial *Legionella* test-negative pneumonia were enrolled. Both cohorts were representative of their respective condition. Key differences between Legionnaires’ disease and non-*Legionella* bacterial pneumonia patient groups emerged in terms of comorbidities, pneumonia severity, and self-reported quality of life. These differences will be accounted for in future analyses as part of the *LongLEGIO* study.

**Conclusions:** The *LongLEGIO* study will advance ongoing research on post-acute infection syndromes and provide a robust data foundation for more accurate assessments of the disease burden associated with Legionnaires’ disease.

## Background

Lower respiratory tract infections, including pneumonia, remain a major cause of morbidity and mortality worldwide [1, 2] and pose a significant burden on healthcare systems [3, 4]. Manifestations of pneumonia can furthermore persist for weeks to months after the acute infection and may lead to additional use of health service [5–7]. Persistent sequelae of pneumonia include respiratory symptoms such as cough or dyspnoea [8], cardiovascular complications [9], cognitive impairment [10–12], general fatigue [8, 13, 14], and reduced quality of life [15]. Moreover, extrapulmonary sequelae often outlast respiratory symptoms [5, 13, 16]. The extent to which such persistent sequelae of pneumonia are influenced by the infection-causing pathogens is poorly understood. This is because most studies either do not differentiate between different pneumonia-causing pathogens in the analysis or only stratify for *Streptococcus pneumonia*, the most frequently identified cause of community-acquired pneumonia (CAP) [13, 17].

To better understand the impact of specific pathogens on persistent pneumonia manifestations, Legionnaires’ disease (LD) is particularly interesting to study. LD is caused by gram-negative *Legionella* bacteria and accounts for approximately 4-7% of CAP cases in Europe [18]. In addition to respiratory symptoms, LD is often associated with extrapulmonary manifestations such as confusion, diarrhoea, headache and acute kidney damage [19–21]. The underlying pathophysiological mechanisms of *Legionella* infections further closely resemble those of *Coxiella burnetii* infections causing Q-fever [22, 23]. The long-term health effects of Q-fever, particularly chronic fatigue, are well documented in the literature [24].

In contrast to Q-fever, persistent health impacts of LD remain poorly understood, and studies on this topic are scarce. Lettinga *et al.* reported that LD patients experienced persistent fatigue, neurological symptoms such as concentration difficulties and memory loss, muscle weakness, and reduced quality of life up to 17 months after acute infection [25]. However, it remained unclear whether these persistent health impairments were due to the *Legionella* infection or the severity of the pneumonia. Similarly, another cross-sectional study compared Q-fever and LD patients with healthy subjects from the general population one year after the acute infection. This study also suggested that LD, like Q-fever, can lead to a persistent reduction in quality of life and chronic fatigue [26]. By design, however, the study did not allow to draw conclusions about the temporal relationship between infection and the presence of the observed health impairments. To date, no study has prospectively compared the persistent health impacts of community-acquired LD (CALD) with the lasting health effects of other bacterial CAP or systematically assessed the use of health services by CALD patients beyond the acute phase of infection.

Here, we present the *LongLEGIO* study and report on the study enrolment. The study explores the persisting impacts of CALD on patients’ health, well-being, and health service use, and compares LD patients with other bacterial CAP patients tested negative for a *Legionella* infection.

## Methods

### Study design and objectives

The *LongLEGIO* study is a matched, prospective observational cohort study. The study enrolled community-acquired Legionnaires’ disease (CALD) and *Legionella* test-negative bacterial CAP (non-LD bacterial CAP) patients from secondary and tertiary care hospitals in Switzerland.

The *LongLEGIO* study addresses the following objectives: (i) to explore CALD patients’ symptoms and general well-being and compare them to symptoms and the well-being of other non-LD bacterial CAP patients, (ii) to describe and compare the care needs and health service utilisation of CALD and non-LD bacterial CAP patients during the recovery, including their challenges in navigating the health system.

### Recruitment and data collection

For the recruitment and data collection, *LongLEGIO* builds on an ongoing Swiss national case-control and molecular source attribution study investigating risk factors and infection sources for CALD in Switzerland (*SwissLEGIO*) [27]. The *SwissLEGIO* study recruited 204 CALD patients from 20 secondary- and tertiary-care hospitals across Switzerland between August 2021 and March 2024. The 20 university- and cantonal hospitals jointly reported about 45% of all CALD cases that were notified to the Swiss national notification system for infectious diseases during the recruitment period.

The *LongLEGIO* study capitalises on the *SwissLEGIO* parent study in two ways (**Figure 1, grey boxes**): First, the *LongLEGIO* study recruited CALD patients from a pool of LD patients who previously participated in *SwissLEGIO*. Secondly, the baseline questionnaire and the data extraction from electronic medical records for CALD patients were done as part of the parent study (**Figure 1**).

**Figure 1:**
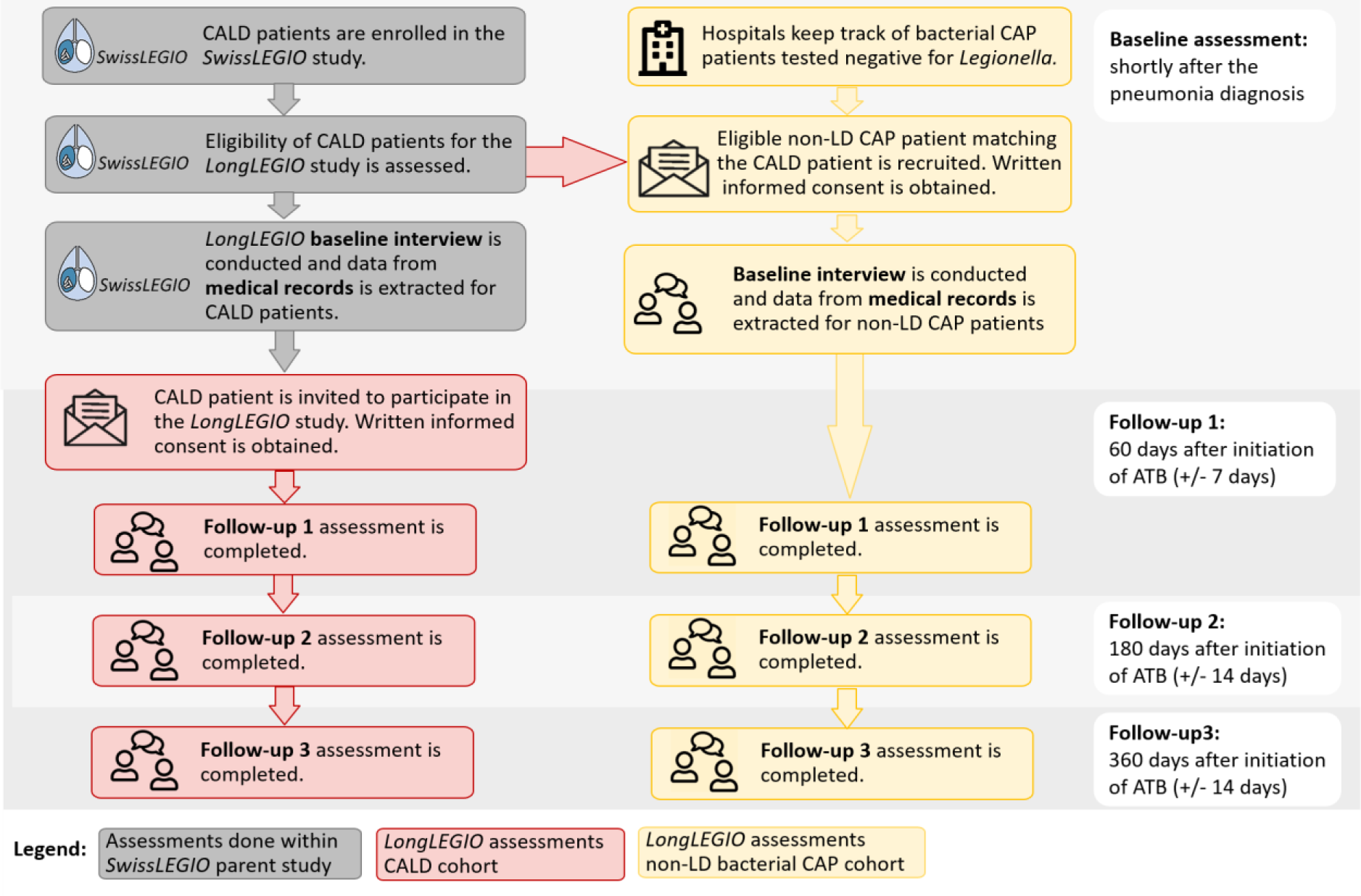
Recruitment, enrolment and data collection for the community-acquired Legionnaires’ disease (CALD) and Legionella test-negative bacterial CAP (non-LD bacterial CAP) cohorts. Grey: Baseline interviews and the extraction of data from electronic medical records for CALD patients is done within the context of the SwissLEGIO parent study. Red: Recruitment and data collection for the CALD cohort. Yellow: Recruitment and data collection for the non-LD bacterial CAP cohort. ABT: appropriate antibiotic therapy.

### Recruitment of community-acquired Legionnaires’ disease (CALD) patients

CALD patients, who previously participated in the *SwissLEGIO* study, were eligible to participate in the *LongLEGIO* study if they fulfilled the study criteria summarised in **Table 1**. Six weeks after CALD patients started appropriate antibiotic therapy^1^, they were invited to participate in the *LongLEGIO* study by postal mail or e-mail. Written informed consent was obtained for all CALD patients prior to the two-month follow-up interview (**Figure 1**).

**Table 1:** Eligibility criteria for CALD patients and non-LD bacterial CAP patients’ participation in the LongLEGIO study.

|  | Inclusion criteria | Exclusion criteria |
| --- | --- | --- |
| **CALD** | – living in Switzerland and speaking German, French, Italian or English<br>– age $\geq 18$ years<br>– health status allows the provision of informed consent and participation in the study<br>– clinical signs and symptoms suggestive of Legionnaires' disease (confirmed pneumonia*)<br>– laboratory confirmed *Legionella* spp. infection (Urinary Antigen Test, PCR or culture) ** | – suspected hospital-acquired pneumonia<br>– suspected travel-associated pneumonia<br>– no COVID-19 laboratory test was performed**<br>– laboratory confirmed COVID-19 infection at hospital admission |
| **Non-LD CAP** | – living in Switzerland and speaking German, French, Italian or English<br>– age $\geq 18$ years<br>– health status allows the provision of informed consent and participation in the study<br>– confirmed pneumonia*<br>– confirmed or suspected bacterial origin of pneumonia***<br>– laboratory test for *Legionella* spp. (Urinary Antigen Test, PCR or culture) performed and negative** | – suspected hospital-acquired pneumonia<br>– suspected travel-associated pneumonia<br>– no COVID-19 laboratory test was performed**<br>– laboratory confirmed COVID-19 infection at hospital admission |

\* Confirmed pneumonia was defined as the presence of a new infiltrate (chest X-ray, ultrasound; or CT scan) PLUS clinical symptoms suggestive of pneumonia (fever, chills, cough, sputum, dyspnoea, tachypnoea, thoracic pain).
\*\* *Legionella* Urinary Antigen Test, PCR or culture and COVID-19 laboratory test were done as part of the routine diagnostics and clinical case management of the patient at the hospital level
\*\*\* Pneumonia must be classified as (suspected) bacterial in the medical records and treated with a full course of antibiotics (defined as empiric or targeted treatment for bacterial pneumonia according to Swiss National Pneumonia guidelines and for at least 3 days [29]. Viral-bacterial pneumonia with bacterial superinfections were considered when treated with a full course of antibiotics.

### Recruitment of *Legionella* test-negative bacterial CAP (non-LD bacterial CAP) patients

*Legionella* test-negative bacterial CAP (non-LD bacterial CAP) patients were recruited through three cantonal (secondary hospitals) and one university hospital (tertiary hospital). All three hospitals also participate in the *SwissLEGIO* parent study. During the recruitment period, all hospitals kept track of all bacterial CAP patients who tested negative for *Legionella* and COVID-19 and who were treated with a full course of antibiotics for pneumonia.^2^ The recruitment of non-LD bacterial CAP patients occurred shortly after the patients were diagnosed with pneumonia (**Figure 1**).

Non-LD bacterial CAP patients were matched to CALD patients based on sex, age (+/- 5 years), hospital level (university vs. cantonal hospitals), and date of diagnosis (+ 60 days). The recruitment of the non-LD bacterial CAP patients was triggered by the enrolment of a CALD patient – eligible for the *LongLEGIO* study – into the *SwissLEGIO* study. In short, the study team sent a recruitment request with the matching criteria to one of the partner hospitals. The hospital then recruited the eligible non-LD bacterial CAP patient with the closest date of a matching diagnosis. Eligibility criteria for non-LD bacterial CAP patients are summarised in **Table 1**. Written informed consent was obtained prior to the baseline interview.

### Data collection

For the *LongLEGIO* study, the two cohorts of CALD patients and non-LD bacterial CAP patients are followed up over 12 months. Assessments are done at four different time points **(Figure 1):** Shortly after the pneumonia diagnosis, a semi-structured baseline interview is conducted and additional data is extracted from electronic medical records for the current hospitalisation. Follow-up assessments in the form of semi-structured interviews are afterwards conducted at two, six and twelve months after patients started appropriate antibiotic therapy^3-4^. All interviews are either conducted in person or over the phone/ by video calls.

At baseline, the questionnaire consists of structured and open questions related to patients’ pre-existing co-morbidities, their acute illness experience, the perceived disease severity and patients’ health-seeking (**Table 2, Questionnaire**). In addition to the baseline interview, information on the patient’s medical history, disease severity, information on the pneumonia causing pathogen, the clinical case management and prescribed antibiotics were extracted from electronic medical records.

**Table 2:** Content of the LongLEGIO questionnaire.

| **Baseline questionnaire** |  |
| --- | --- |
| Patient characteristics | – Demographic information (e.g. sex, age, education, income) |
| Illness experience and health-seeking for acute pneumonia | – Pneumonia manifestation<br>– Disease perception and perceived severity of the pneumonia<br>– Health-seeking prior to hospital admission |
| Comorbidities and health-related quality of life (HRQoL) | – Comorbidities<br>– HRQoL prior to pneumonia (EQ-5D-5L) [30] |
| **Follow-up questionnaire** |  |
| Perceived recovery from pneumonia and health related quality of life (HRQoL) | – Subjective judgment on overall health<br>– Subjective assessment of the degree of recovery from pneumonia<br>– 5-item World Health Organisation Well-being Index (WHO-5) [31]<br>– HRQoL at time of follow-up interview (EQ-5D-5L) [30] |
| Persistent symptoms | – Presence and perceived severity of symptoms (extensive list of pneumonia symptoms reported in the literature is provided). Patients are asked to judge if symptoms are related to the pneumonia.<br>– Quantification of dyspnoea (modified Medical Research Council, mMCR) [32]<br>– Fatigue assessment scale (FAS) [33]<br>– Worsening of pre-existing comorbidities or newly diagnosed medical conditions |
| Health care utilisation and patients' health seeking behaviour | – Quantification of formal healthcare utilization since last assessment<br>– Quantification of health seeking outside the formal health sector since last assessment<br>– Changes in prescribed/ newly prescribed medications<br>– Patient experiences in navigating the health system and perceived challenges |
| Social life and work participation | – If persisting symptoms: impact on work participation and social life |

The follow-up questionnaires consist of structured and open questions to investigate (i) the patient’s perceived health, recovery, and health-related quality of life after the acute pneumonia, (ii) the presence of persistent pneumonia-related symptoms, (ii) patients’ (potentially extended) health service utilisation and health seeking in the informal health sector after the hospital discharge, and (iv) potential impacts of the infection on patients’ social and work life (**Table 2**). The design of the follow-up questionnaires was informed by the Explanatory Model Interview Catalogue (EMIC) [34], by research on the long-term recovery from pneumonia [8, 15, 35–37] and by research on Long-COVID [38–40].

### Statistical analysis

CALD patients and non-LD bacterial CAP patients are characterised in terms of demographics, illness experience, help-seeking, comorbidities and health-related quality of life (HRQoL). Continuous variables are presented as medians (IQRs), and categorical variables as n (%).

To compare persistent symptoms, additional healthcare consultations after discharge, and changes in HRQoL scores over time (two, six and 12 months) within each cohort, we will be use the McNemar’s chi-square test or the Wilcoxon signed-rank test as appropriate. For comparisons between groups at each time point, we will use the Chi-square test or Fisher’s exact test for categorical variables, and the t-test or Mann-Whitney U test for continuous variables, as appropriate.

The association between CALD and long-term symptoms will be evaluated by multivariable logistic regression, adjusting for potential confounders such as age, gender, smoking status, comorbidities, and severity of pneumonia. Given the large proportion of elderly participants, a subgroup analysis will be conducted for patients aged 65 and older.

Only participants who completed at least the first follow-up interview will be included in the analysis. If possible, multiple imputation for missing variables and a sensitivity analysis using complete-case analysis is foreseen.

Statistical analysis will be conducted using the statistical software R Version 4.3.2 (R Core Team, Vienna, Austria).

### Data management

Data are collected on standardised electronic Case Report Form (eCRF) using the data collection software Open Data Kit (ODK, getodk.org). Forms are encrypted and all patient-identifying information is removed. Individual forms are linked through unique subject IDs. Automated validations are implemented in the eCRF to check for data completeness and plausibility. In addition, submitted forms are continuously checked for plausibility and accuracy, and source data verification is performed. Data is stored on a secured network drive accessible only to authorised study personnel. The network drive is backed up regularly, according to our internal institutional policy.

## Study population

### Sample size and enrolment

The recruitment timelines and the sample size for the *LongLEGIO* study were bound to the enrolment of CALD patients in the *SwissLEGIO* parent study. Overall, 86 CALD patients and 110 non-LD bacterial CAP patients were invited to participate in the *LongLEGIO* study between June 2023 and June 2024. In total, 59 CALD patients (enrolment rate of 69%) and 60 non-LD bacterial CAP patients (enrolment rate of 55%) agreed to participate. There were no differences in age, gender and ICU admission rates between participants and non-participants among CALD patients, and in gender and ICU admission rates among the patients with non-LD bacterial CAP (**Table S1, Supplementary material**). There was a statistically significant difference in age between participants and non-participants among the non-LD bacterial CAP patients (69 vs. 77, p=0.016).

With 59 CALD patients and 60 non-LD bacterial CAP patients enrolled in the *LongLEGIO* study, the sample size was sufficient to provide a power of 78% with alpha=0.05 to detect a 20% difference between our two cohorts. Given that the majority of patients in both groups were over 50 years old, we assumed for our power calculation a 10% prevalence of non-specific symptoms—such as fatigue, weakness, and a general reduction in quality of life—that were not clearly attributable to either CALD or bacterial CAP.

### Baseline characteristics

The baseline characteristics for the CALD and the non-LD bacterial CAP cohorts are summarised in Table 3. The median age for both cohorts is 69 years; males comprised 59.3% among CALD and 63.3% among non-LD bacterial CAP patients. Both cohorts have a similar comorbidity burden as measured by the Charlson Comorbidity Index (CCI). Non-LD bacterial CAP patients, however, were more likely to suffer from chronic obstructive pulmonary disease (20% vs 10.2%), malignancies (31.7% vs 13.6%), and were more likely immunosuppressed (23.3% vs 13.6%). On the contrary, more CALD than non-LD bacterial CAP patients suffered from chronic kidney failure (15.3% vs 8.3%). Patients in both cohorts exhibited a similar health-seeking behaviour prior to the hospital visit with the majority of patients initially consulting a GP. CALD patients were more frequently pre-treated with antibiotics (28.3% vs 15.3%).

**Table 3:** Baseline characteristics of 119 Legionnaires’ disease patients (CALD) and non-LD bacterial community-acquired pneumonia patients (CAP)

|  | CALD (n=59) | non-LD bacterial CAP (n=60) |
| --- | --- | --- |
| **Patient characteristics** |  |  |
| Male | 35 (59.3 %) | 38 (63.3 %) |
| Age (median [IQR]) | 69.0 [56.5, 79.5] | 69.0 [59.8, 79.0] |
| Income |  |  |
| <60'000 CHF | 15 (36.6%) | 21 (39.6%) |
| 60'001-100'000 CHF | 21 (51.2%) | 22 (41.5%) |
| >100'000 CHF | 5 (12.2%) | 10 (18.9%) |
| Body Mass Index (BMI) [kg/m2] (median [IQR]) | 24.0 [22.0, 28.0] | 25.0 [21.0, 28.0] |
| **Medical conditions** |  |  |
| Comorbidities<sup>1</sup> |  |  |
| Heart disease | 21 (35.6 %) | 22 (36.7 %) |
| Heart failure | 7 (11.9 %) | 8 (13.3 %) |
| Diabetes mellitus | 7 (11.9 %) | 10 (16.7 %) |
| COPD | 6 (10.2 %) | 12 (20 %) |
| Chronic kidney failure | 9 (15.3 %) | 5 (8.3 %) |
| Cerebrovascular diseases | 5 (8.5 %) | 6 (10 %) |
| Malignancies (haematological/solid organ) | 8 (13.6 %) | 19 (31.7 %) |
| Immunosuppression | 8 (13.6 %) | 14 (23.3 %) |
| Charlson Comorbidity Index (CCI) (median [IQR]) | 1.00 [0.00, 2.00] | 1.00 [0.00, 3.00] |
| Active smokers (self-reported) | 20 (33.9 %) | 13 (22.4 %) |
| **Health seeking prior to hospital visit** |  |  |
| Medical advice prior to admission |  |  |
| None | 20 (34.5 %) | 18 (30.5 %) |
| General practitioner | 28 (48.3 %) | 26 (44.1 %) |
| Hospital/Emergency department | 2 (3.4 %) | 6 (10.2 %) |
| Specialist doctor | 4 (6.9 %) | 5 (8.5 %) |
| Care giver/Family members/Close friends | 7 (12.1 %) | 6 (10.2 %) |
| Antibiotics prescribed before current pneumonia / hospital admission |  |  |
| None | 39 (66.1) | 50 (83.3) |
| Co-Amoxicillin | 15 (25.4) | 6 (10.0) |
| Cephalosporins | 1 ( 1.7) | 0 ( 0.0) |
| Combination therapies<sup>2</sup> | 1 ( 1.7) | 1 ( 1.7) |
| Prior antibiotic treatment with unknown agent | 3 ( 5.1) | 3 ( 5.0) |
| Time from symptom onset to hospital admission, days (median [IQR]) | 4.0 [2.0, 6.0] | 5.0 [2.8, 10.0] |
| **Self-reported symptoms during acute pneumonia episode** |  |  |
| Fatigue/Weakness | 55 (94.8 %) | 49 (83.1 %) |
| Fever | 50 (89.3 %) | 45 (76.3 %) |
| Cough | 38 (66.7 %) | 48 (80.0 %) |
| Shortness of breath | 37 (66.1 %) | 46 (78.0 %) |
| Muscle aches | 29 (51.8 %) | 15 (25.9 %) |
| Headache | 26 (47.3 %) | 16 (28.1 %) |
| Chest pain | 20 (35.1 %) | 28 (49.1 %) |
| Confusion | 19 (33.3 %) | 20 (35.1 %) |
| Diarrhoea | 15 (26.3 %) | 17 (28.8 %) |
| Nausea/Emesis | 7 (11.9 %) | 2 (3.3 %) |
| Loss of appetite | 5 (8.5 %) | 4 (6.7 %) |
| Self-rate severity from scale 1 to 10 (median [IQR]) | 8.0 [7.0, 9.0] | 7.0 [5.0, 8.0] |
| **Progression of CAP and severity** |  |  |
| Time from hospital admission to start of adequate antibiotic treatment, days (median [IQR]) | 0.0 [0.0, 1.0] | 0.00 [0.0, 1.0] |
| Length of hospital stay, days (median [IQR]) | 6.0 [4.0, 8.5] | 6.0 [4.0, 7.0] |
| ICU admission | 8 (13.6 %) | 5 (8.3 %) |
| Non-invasive ventilation | 7 (11.9 %) | 12 (28.6 %) |
| Invasive ventilation | 4 (6.8 %) | 1 (1.8 %) |
| Discharge followed by |  |  |
| Ambulatory care | 47 (81.0 %) | 52 (86.7 %) |
| Referral to another hospital | 5 (8.6 %) | 0 (0.0 %) |
| Referral to a rehabilitation centre | 6 (10.3 %) | 8 (13.3 %) |
| **Quality of life (EQ-5D-5L)<sup>3</sup>** |  |  |
| Mobility | 14 (23.7 %) | 22 (36.7 %) |
| Self-care | 2 (3.4 %) | 3 (5.0 %) |
| Usual activities | 13 (22.0 %) | 20 (33.3 %) |
| Pain/Discomfort | 20 (34.5 %) | 26 (43.3 %) |
| Anxiety/Depression | 20 (35.1 %) | 20 (33.3 %) |
| EQ-VAS score (median [IQR]) | 85.0 [75.0, 90.0] | 70.0 [50.0, 85.0] |

<sup>1</sup> as reported in the electronic medical records
<sup>2</sup> Co-Amoxicillin or Cefepim in combination with macrolides
<sup>3</sup> The five dimensions of the EQ-5D-5L quality of life assessment were dichotomised: 0 indicates no problem, 1 indicates any problem from mild to severe (details for each dimensions are provided in the Supplementary material)

During the acute phase of their pneumonia, non-LD bacterial CAP patients more frequently reported respiratory symptoms including cough (80% vs 66.7%), shortness of breath (78% vs 66.1%), and chest pain (49.1% vs 35.1%). On the contrary, CALD patients reported more frequently extrapulmonary symptoms such as fever (89.3% vs 76.3%), muscle aches (51.8% vs 25.9%), headaches (47.3% vs 28.1%) and nausea or emesis (11.9% vs 3.3%). The ICU admission rate and the proportion of patients who needed mechanical ventilation was higher for CALD than for non-LD bacterial CAP patients (13.6% vs 8.3% and 6.8% vs 1.8%, respectively).

Finally, participants were asked to report on their perceived quality of life prior to experiencing first pneumonia symptoms (**Table S2, Supplementary material**). Generally; non-LD bacterial CAP patients reported lower quality of life scores compared to CALD patients. Differences were particularly pronounced for the EQ-5D-5L dimensions of mobility, daily activity and pain/discomfort.

## Discussion

Persistent manifestations of CALD beyond the acute infection and its impact on healthcare utilisation remain poorly understood. We present the rationale and study design of a prospective, matched cohort study to investigate persistent sequelae of CALD and compare them to manifestations of persistent pneumonia among non-LD bacterial CAP patients.

Previous studies examining the persistent sequelae of CAP have rarely stratified their analysis by the different pneumonia-causing pathogens [9–12, 14, 15]. Among those that have, the focus was on *Streptococcus pneumonia* [13, 17]. This limited stratification was mainly due to sample size limitations and lack of information on the CAP-causing pathogen in these studies. In contrast, studies focusing specifically on persistent sequelae of *Legionella* infections never included pneumonia patients as a comparison group. To our knowledge, the *LongLEGIO* study is the first to systematically compare the persistent health effects of CALD with the persistent manifestations of non-CALD bacterial CAP, going beyond a survival analysis based on mortality data [41]. Such a comparison is crucial to contextualise the frequency of observed health and well-being impairments in CALD patients against the background occurrence in a suitable control group. It will further allow us to investigate the extent to which the persistence of health impairments of CAP is reflected in the tropism and pathogenic mechanisms of the causative pathogen. Ultimately, this may improve our understanding of the underlying pathogenesis of persistent pneumonia manifestations and post-acute infection syndromes [24, 42].

The *LongLEGIO* study focuses on patient-reported outcome measures (PROMS), as opposed to selecting mortality or hospital readmission rates as primary outcome measures, which has been the focus of previous studies on *Legionella* [41, 43]. PROMS provide robust measures of persistent symptoms and functional impairments that patients experience [8]. Persistent symptoms and functional impairments, as measured by PROMS, are also one of the main reasons for additional GP consultations and emergency department visits after the acute phase of pneumonia [6, 7]. PROMS, however, are usually not well documented in medical records and hence can only be adequately captured by applying prospective study designs [42].

The *LongLEGIO* study is also the first study to assess the healthcare utilisation of CALD patients beyond the acute infection and to explore its association with persistent sequelae of CALD. As the COVID-19 pandemic and the advent of Long COVID have clearly demonstrated, post-acute infection syndromes can contribute significantly to the disease burden of infectious diseases [39, 42]. However, to date, estimates of the burden of LD do not take into account persistent health impairment and the associated increased medical expenditure [44, 45]. By providing insights into both, the presence of persistent health and well-being impairment and additional healthcare utilisation after the acute infection, the *LongLEGIO* study will generate data to facilitate a better estimation of the disease burden of LD in the future. This, in turn, may help public health specialists and physicians to better plan and anticipate the health care resources needed for the holistic treatment of CALD.

Enrolment for the *LongLEGIO* study was completed in June 2024. A total of 59 CALD patients and 60 non-LD bacterial CAP patients have been enrolled. Our non-LD bacterial CAP and CALD patient cohorts are representative for the two patient populations. Similar to previous studies comparing characteristics of CALD and non-LD bacterial CAP patients, LD patients were more likely to suffer from extrapulmonary symptoms, were more frequently pre-treated with antibiotics prior to hospital admission and were more often admitted to the ICU [21, 46, 47]. Non-LD bacterial CAP patients, on the other hand, were more likely to suffer from chronic obstructive pulmonary disease and malignancies, which is also consistent with findings from previous research [20]. Finally, we also observed differences in the self-reported quality of life scores (lower scores were reported by non-LD bacterial CAP patients). Both, comorbidities and pneumonia severity may influence the long-term health outcomes after pneumonia [5, 6, 13, 16]. We will, therefore, correct for these differences in our analyses. For the self-reported quality of life score, each patient will serve as their own control and relative rather than absolute changes in reported scores from the pre-pneumonia baseline score will be assessed.

The *LongLEGIO* study has limitations. Despite its prospective nature, the study may be subject to recall bias, especially between the six- and 12-month follow-up periods. We try to mitigate this recall bias by specifically probing for both, intermittent and current symptoms. In addition, interviewers are instructed to systematically probe for all the symptoms and health service consultations that were reported by the patient in previous interviews. Finally, and despite the integration of the *LongEGIO* study into the national *SwissLEGIO* parent study, our sample size for CALD remains relatively small. It is, therefore, possible that we underestimate functional impairments that are relatively rare although we are confident to fully capture the common persistent sequelae of CALD.

In summary, the *LongLEGIO* study explores and compares persistent health and well-being impacts of CALD and non-LD bacterial CAP in two representative cohorts. The study aims to contribute to ongoing research on post-acute infection syndromes, a phenomenon long recognized and well-documented for conditions like Q-fever but often challenging to capture in daily medical practice [24]. As the recent COVID pandemic has shown, more research on persistent health impairments after acute infections is highly needed as they represent a significant burden for patients and healthcare systems.

## Data Availability

All data produced in the present work are contained in the manuscript.

## Ethical considerations

Ethical approval for the study was obtained from the Ethics Commission of Northwestern and Central Switzerland (EKNZ, 2023-00639). This study is conducted in accordance with the principles of Good Epidemiological Practice and the Declaration of Helsinki. Data are stored in concordance with Swiss data protection laws. Written informed consent was obtained from all study participants prior to enrolment.

## Acknowledgements

We thank Jan Hattendorf for his valuable advice on the design and development of the protocol. At the Federal Office of Public Health, we acknowledge the various initial inputs and exchanges surrounding this study with Ornella Luminati, Monica Wymann and Mirjam Mäusezahl-Feuz. We thank Chiara Strozzi and the Kompetenzzentrum für Ausbruchs-untersuchungen (KEA) at Swiss TPH for their support in the final data collection phase.

## Supplementary material

**Table S1:** Baseline characteristics of 86 Legionnaires’ disease patients (CALD) and 110 non-LD bacterial community-acquired pneumonia patients (CAP) invited to participate in the LongLEGIO study between June 2023 and June 2024.

|  | Enrolled | Not enrolled | p value |
| --- | --- | --- | --- |
| **CALD patients** | **N=59** | **N=27** |  |
| Age (median [IQR]) | 69.0 [56.6, 79.5] | 65.0 [59.5, 75.0] | 0.589 |
| Male | 35 (59.3%) | 17 (63.0 %) | 0.934 |
| Hospitalisation | 57 (96.6 %) | 27 (100.0 %) | 0.844 |
| ICU admission | 8 (13.6 %) | 4 (14.8 %) | 1.000 |
| **Non-LD bacterial CAP** | **N=60** | **N=50** |  |
| Age (median [IQR]) | 69.0 [59.8, 79.0] | 77.0 [67.0, 82.0] | 0.016 |
| Male | 22 (36.7 %) | 19 (38.0 %) | 1.000 |
| Hospitalisation | 58 (96.7 %) | 43 (100.0 %) | 0.628 |
| ICU admission | 5 (8.3 %) | 1 (2.4 %) | 0.423 |

**Table S2:** Supplementary baseline characteristics of 119 Legionnaires’ disease patients (CALD) and non-LD bacterial community-acquired pneumonia patients (CAP)

|  |  | CALD (n=59) | non-LD bacterial CAP (n=60) |
| --- | --- | --- | --- |
| **Quality of Life (EQ-5D-5L)** |  |  |  |
| Mobility | No problem | 45 (76.3 %) | 38 (63.3 %) |
|  | Slight problem | 5 (8.5 %) | 8 (13.3 %) |
|  | Moderate problem | 6 (10.2 %) | 10 (16.7 %) |
|  | Severe problem | 3 (5.1 %) | 4 (6.7 %) |
|  | Unable | 0 (0.0 %) | 0 (0.0 %) |
| Self-care | No problem | 57 (96.6 %) | 57 (95.0 %) |
|  | Slight problem | 2 (3.4 %) | 3 (5.0 %) |
|  | Moderate problem | 0 (0.0 %) | 0 (0.0 %) |
|  | Severe problem | 0 (0.0 %) | 0 (0.0 %) |
|  | Unable | 0 (0.0 %) | 0 (0.0 %) |
| Usual activities | No problem | 46 (78.0 %) | 40 (66.7 %) |
|  | Slight problem | 7 (11.9 %) | 13 (21.7 %) |
|  | Moderate problem | 3 (5.1 %) | 5 (8.3 %) |
|  | Severe problem | 3 (5.1 %) | 2 (3.3 %) |
|  | Unable | 0 (0.0 %) | 0 (0.0 %) |
| Pain/Discomfort | No problem | 37 (64.9 %) | 34 (56.7 %) |
|  | Slight problem | 13 (22.8 %) | 5 (8.3 %) |
|  | Moderate problem | 5 (8.8 %) | 15 (25.0 %) |
|  | Severe problem | 2 (3.5 %) | 6 (10.0 %) |
|  | Unable | 0 (0.0 %) | 0 (0.0 %) |
| Anxiety/Depression | No problem | 37 (64.9 %) | 40 (66.7 %) |
|  | Slight problem | 13 (22.8 %) | 11 (18.3 %) |
|  | Moderate problem | 5 (8.8 %) | 5 (8.3 %) |
|  | Severe problem | 2 (3.5 %) | 4 (6.7 %) |
|  | Unable | 0 (0.0 %) | 0 (0.0 %) |
| **Usual health-seeking prior to infection** |  |  |  |
| General practitioner |  | 49 (83.1 %) | 46 (78.0 %) |
| Hospital/Emergency department |  | 4 (6.8 %) | 4 (6.7 %) |
| Specialist doctor |  | 5 (8.5 %) | 4 (6.7 %) |
| Care giver/Family members/Close friends |  | 18 (30.5 %) | 10 (16.7 %) |
| Pharmacy |  | 10 (16.9 %) | 1 (1.7 %) |
| Telemedicine |  | 2 (3.4 %) | 2 (3.3 %) |

|  | CALD (n=59) | non-LD bacterial CAP (n=60) |
| --- | --- | --- |
| **Self-reported pre-existing comorbidities** |  |  |
| High blood pressure<sup>1</sup> | 22 (37.9 %) | 24 (41.4 %) |
| Ischemic heart disease | 7 (11.9 %) | 11 (18.3 %) |
| Heart failure | 3 (5.3 %) | 7 (12.1 %) |
| Cerebrovascular disease in the past | 4 (6.8 %) | 8 (13.8 %) |
| COPD | 6 (10.3 %) | 5 (8.6 %) |
| Asthma | 8 (13.8 %) | 10 (17.5 %) |
| Pneumonia in the last 5 years | 7 (12.1 %) | 12 (21.1 %) |
| Cancer (ongoing + in the last 5 years) | 7 (11.9 %) | 15 (25.4 %) |
| Diabetes mellitus | 7 (11.9 %) | 7 (12.1 %) |
| Immunosuppression | 9 (15.8 %) | 14 (26.4 %) |
| Chronic liver disease | 1 (1.7 %) | 5 (8.5 %) |
| Chronic kidney disease | 4 (7.0 %) | 8 (14.0 %) |
| **Aetiology of non-LD pneumonia** |  |  |
| Unspecified aetiology |  | 44 (73.3 %) |
| Streptococcus pneumoniae |  | 5 (8.3 %) |
| Streptococcus pneumoniae and Haemophilus influenzae |  | 1 (1.7 %) |
| Mycoplasma pneumoniae |  | 3 (5.0 %) |
| Haemophilus influenzae |  | 3 (5.0 %) |
| Haemophilus influenzae and Rhinovirus pneumoniae |  | 2 (3.3 %) |
| Staphylococcus aureus and Klebsiella pneumoniae |  | 1 (1.7 %) |
| Pseudomonas aeruginosa |  | 1 (1.7 %) |

<sup>1</sup>Based on clinical evaluation

## Footnotes

1 Defined as the prescription of macrolides, quinolones or doxycycline. All these antibiotics exhibit intracellular activity and hence, activity against *Legionella*. [22, 28]

2 Defined as empiric or targeted treatment for bacterial pneumonia according to Swiss National Pneumonia guidelines and for at least 3 days. [29]

3 For CALD patients, this was defined as the prescription of macrolides, quinolones or doxycycline. [22, 28]

4 For non-LD bacterial CAP patients, this was defined as the prescription of beta-lactams, macrolides, quinolones or tetracyclines for patients with pneumonia of unclear aetiology OR pathogen-specific antibiotics if aetiology was known. [29]

